# *SYNE1* gene novel variant is associated with myocardial infarction in young people with a family history of premature atherosclerosis

**DOI:** 10.1101/2024.09.06.24313220

**Authors:** Michał Ambroziak, Jakub Franke, Anna Wójcicka, Monika Kolanowska, Andrzej Budaj

## Abstract

**Aim:** The aim of the study was to investigate the role of genetic variants in young patients (aged <50 years) with myocardial infarction (MI) and a family history of premature atherosclerosis.

**Methods and Results:** The studied group consisted of 70 patients aged 26-49 (mean 43.1, SD ±4.3), 17 women and 53 men, with MI and with a family history of premature atherosclerosis, defined as MI or ischaemic stroke in first-degree relatives at age <65 years in women or <55 years in men. The total DNA was extracted from the peripheral blood samples. The targeted enrichment library was prepared and analyzed using the Next-Generation Sequencing method. Statistical analyses were performed using the R software package (http://www.r-project.org/). The results of sequencing were compared to data from the reference control population consisting of 597 people with no history of MI (418 women, 179 men) aged 18-83 (mean 40.5, SD ± 12.4) as a whole and after matching with a studied group by age and gender in a proportion 1:3 (210 people, 51 women, 159 men, aged 18-77, mean 42.1, SD ±10.6) using Propensity Score Matching. Risks associated with detected variants were evaluated using Fisher’s exact test based on the allelic frequencies of variants in both groups.

*SYNE1* gene variant rs36215567 (NM_182961.4: c.20396+22A>G) occurs with a significantly higher incidence in the studied group when compared to the control population with OR 4.80 95%CI 1.43-14.45 (p=0.005) as well as when compared to the control population matched by age and gender OR 9.31 95%CI 1.64-95.41 (p=0.004). There were no statistically significant differences in the incidence of variants related to familial hypercholesterolemia such as *LDLR* c.667G>A, *PCSK9* c.658-36G>A, and *APOB* c.12382G>A between both cohorts.

**Conclusion:** A novel variant of the *SYNE1* gene is associated with myocardial infarction in young patients with a family history of premature atherosclerosis.

**Lay summary:**

- The wide use of genetic information to predict coronary artery disease (CAD) development and incidence, particularly important in young people, still requires investigation before clinical implementation.
- A novel variant of the *SYNE1* gene is associated with myocardial infarction (MI) in young patients with a family history of premature atherosclerosis.
- Although, the role of SYNE1 in the maintenance of proper function of the cardiac muscles and coronary arteries remains still under examination the result of this study open a new opportunity to use the *SYNE1* gene variant in genetic risks MI scores.

## 1. Introduction

Cardiovascular diseases (CVDs) are the leading cause of death globally. As the World Health Organization (WHO) reports, an estimated 17.9 million people died from CVDs in 2019, representing 32% of all global deaths [1]. Ischemic heart disease (IHD) rose from the third to the first leading global cause of premature mortality, measured by years of life lost (YLL), between 1990 and 2019 [2]. In the EUROASPIRE V study, which involved people with confirmed coronary artery disease (CAD), up to 9% of the studied population were patients aged <50 years [3]. According to the Framingham study, the MI incidence for a 10-year follow-up differs between the age groups - it was 12.9, 38.2, and 71.2 per 1000 in males and 2.2, 5.2, and 13.0 per 1000 in females in the age groups of 30–34, 35–44, and 45–54 years, respectively [4].

There are differences in risk factor profiles between individuals who experienced the first myocardial infarction at a very young (≤40 years) and a young (age 41-50 years) age. When compared with young patients, very young individuals are more likely to use both marijuana and cocaine [5]. Conversely, patients aged 41 to 50 years are more likely to be diagnosed with hypertension or peripheral vascular disease, use alcohol, and have a higher 10-year atherosclerotic cardiovascular disease risk score. Nevertheless, the overall burden of traditional risk factors was similar between very young and young patients.

The INTERHEART study carried out in 52 countries around the world on more than 12 thousand patients showed that smoking, lipid abnormalities, hypertension, and diabetes as more significant risk factors for MI in younger than in older patients [6]. Apart from those factors, a family history of premature IHD and genetic factors seem to be especially important in young people with MI.

According to the European Society of Cardiology guidelines for CVD prevention, the routine use of genetic risk scores is not yet recommended [7]. There is no consensus regarding which genes and corresponding single nucleotide polymorphisms should be included, and whether to use risk factor-specific or outcome-specific polygenic risk scores (PRS) [8]. The addition of a PRS for CAD to pooled cohort equations seems to be associated with a statistically significant, yet modest, improvement in the predictive accuracy for incidents of CAD and improved risk stratification for only a small proportion of individuals [9]. Among young adults from the CARDIA study (Coronary Artery Risk Development in Young Adults) and Framingham Offspring Study, PRS improved model discrimination for coronary atherosclerosis, but improvements were smaller than those associated with modifiable risk factors [10].

Nevertheless, there is no doubt that genetic information is a valuable complement to traditional factors in CAD risk stratification, particularly in young people. The wide use of genetic information to predict CAD development and incidence still requires investigation before clinical implementation. The group of special interest is a population of young people with CAD and a family history of premature IHD, defined as MI or ischemic stroke in first-degree relatives at age <55 years in men and < 65 years in women [11]. Thus, the aim of our study was to investigate the role of genetic variants in this very special population – young patients with MI and a family history of premature IHD.

## 2. Subjects and methods

### 2.1. Patients

The studied group consisted of 70 patients with MI aged 26-49 (mean 43.1, SD ±4.3; 17 women and 53 men), admitted to the Department of Cardiology, Centre of Postgraduate Medical Education, Grochowski Hospital in years 2007-2019. All patients had been diagnosed MI based on clinical presentation, ecg and biochemical criteria, including MI with ST elevation (STEMI) in 18 cases (25.7%) and MI without ST elevation (NSTEMI) in 52 cases (74.3%) and had a family history of premature atherosclerosis, defined as MI or ischemic stroke in first-degree relatives at age <65 years in women or <55 years in men. All patients, except 2 (no consent), had performed coronary angiography revealing 1-vessel coronary artery disease (VCAD) in 33 patients (47.1%), 2-VCAD in 17 patients (24.3%), and 3-VCAD in 20 patients (28.6%); Table 1. The mean left ventricular ejection fraction (LVEF) was 51.8% (SD ±8.0).

**Table 1.** Clinical and haemodynamic characteristics of studied group (70 patiens). CAD – coronary artery disease, LVEF – left ventricle ejection fraction, NSTEMI – non-ST elevation myocardial infarction, SD – standard deviation, STEMI – ST elevation myocardial infarction.

| Clinical and haemodynamic<br>characteristics | Number of affected patients |
| --- | --- |
|  | n (%) |
| | mean (±SD) |
| **STEMI** | 18 (25.7%) |
| **nSTEMI** | 52 (74.3%) |
| 1-vessel CAD | 33 (47.1%) |
| 2-vessel CAD | 17 (24.3%) |
| 3-vessel CAD | 20 (28.6%) |
| LVEF mean | 51.8% (SD ±8.0) |

There were 15 (21.4%) participants of the study with university degree, 46 (65.7%) with completed high school education and 7 (10%) with completed elementary school education (2 patients have not provided information). Regarding a type of job – there were 37 (52.8%) blue collar workers, 23 (32.9%) white collar workers, 3 (4.3%) unemployed and 2 (2.9%) people on pension (5 patients have not provided information). Regarding marriage status – there were 40 (57.1%) married people in the study, 18 (25.7%) single, 6 (8.6%) divorced, 2 widows and 1widower (4.3%); 3 patients have not provided information.

All participants of the study provided written informed consent. The Ethical Committee of the Centre of Postgraduate Medical Education approved the study protocol. The investigation conforms to the principles outlined in the Declaration of Helsinki.

The results of sequencing were compared to data from the reference Warsaw Genomics control population consisting of 597 people with no history of MI (418 women, 179 men) aged 18-83 (mean 40.5, SD ± 12.4) as a whole and after matching using Propensity Score Matching with a studied group by age and gender in a proportion 1:3 (210 people, 51 women and 159 men, aged 18-77, mean 42.1, SD ±10.6).

### 2.2 Anthropometric, clinical and biochemical analysis

BMI was calculated as weight (kg)/height (m2). Depression and smoking status, including duration and intensity (cigarette number per day) of smoking, were assessed based on the patient’s history. Hypertension was assessed based on the medical history and treatment or on a mean value of two measurements of systolic (SBP) and diastolic (DBP) blood pressure performed after at least 5 minutes sitting, made in 5 minute intervals. Hypertension was defined as values ≥140 mmHg SBP and/or ≥90 mmHg DBP according to ESH/ESC (European Society of Hypertension, European Society of Cardiology) guidelines. Diabetes mellitus was assessed based on the medical history and treatment or based on fasting plasma glucose ≥126 mg/dl or ≥200 mg/dl in oral glucose tolerance test according to WHO and ADA (American Diabetes Association) guidelines.

Blood samples were collected on admission and the next morning. Biochemical analyses, including glucose, total, HDL, and LDL cholesterol as well as triglycerides (TG) plasma concentrations were performed in fasting blood samples by standard enzymatic methods using COBAS INTEGRA 800 regents and equipment (Roche Diagnostics Gmbh).

### 2.3 Genotyping

Total DNA was extracted from whole peripheral blood samples. The extraction was performed on Maxwell® RSC 48 Instrument (Promega) using the Maxwell® RSC Blood DNA Kit, according to the protocol. The targeted enrichment library was prepared and analyzed using the Next-Generation Sequencing (NGS) method. The total DNA input for NGS libraries was 35 ng. The sample libraries were prepared on Biomed i7 (Beckam and Coulter) with the use of Kapa HyperPlus protocol (Roche Diagnostics). The concentration and quality of the libraries were subsequently verified on SpectraMax (Molecular Devices) and TapeStation 4200 and subjected to the hybridization capture (NimbleGen EZ SeqCap, Roche Diagnostics). The hybridization was conducted according to the Roche protocol. The size, concentration, and quality of obtained final NGS library were confirmed by TapeStation 4200 and Quantus (Promega). The samples were sequenced on the Hiseq4000 (Illumina), 2×150 cycles.

### 2.4 Statistical analysis

Statistical analyses were performed using the R software package (http://www.r-project.org/). Risks associated with detected variants were evaluated using Fisher’s exact test based on the allelic frequencies of variants in both groups.

## 3. Results

### 3.1 Anthropometric, biochemical and clinical characteristics of studied and control groups

The mean BMI in the studied group was 28.8 kg/m2 (SD ±3.5), total cholesterol level 202.3 mg/dl (SD ±3.5), HDL 40.6 mg/dl (SD ±9.6), LDL 124.0 mg/dl (SD ±28.9), triglycerides 164.4 mg/dl (SD ±63.5), glucose 108.3 mg/dl (SD ±19.9) and creatinine 0.9 mg/dl (SD ±0.2); Table 2. Regarding other risk factors 63 patients (90%) were smokers (mean 24.1 pack-years, SD ±13.0), 44 (62.8%) had hypertension, 11 (15.7%) had diabetes mellitus, and 8 (11.4%) had depression; Table 3.

**Table 2.** Anthropometric and biochemical characteristics of studied group (70 patients). BMI – body mass index, HDL – high density lipoprotein, LDL – low density lipoprotein, SD – standard deviation.

| **Studied group characteristics** | **mean (±SD)** |
| --- | --- |
| BMI kg/m<sup>2</sup> | 28.8 (±3.5) |
| Total cholesterol mg/dl | 202.3 (±3.5) |
| HDL mg/dl | 40.6 (±9.6) |
| LDL mg/dl | 124.0 (±28.9) |
| Triglycerides mg/dl | 164.4 (±63.5) |
| Glucose mg/dl | 108.3 (±19.9) |
| Creatinine mg/dl | 0.9 (±0.2) |

**Table 3.**
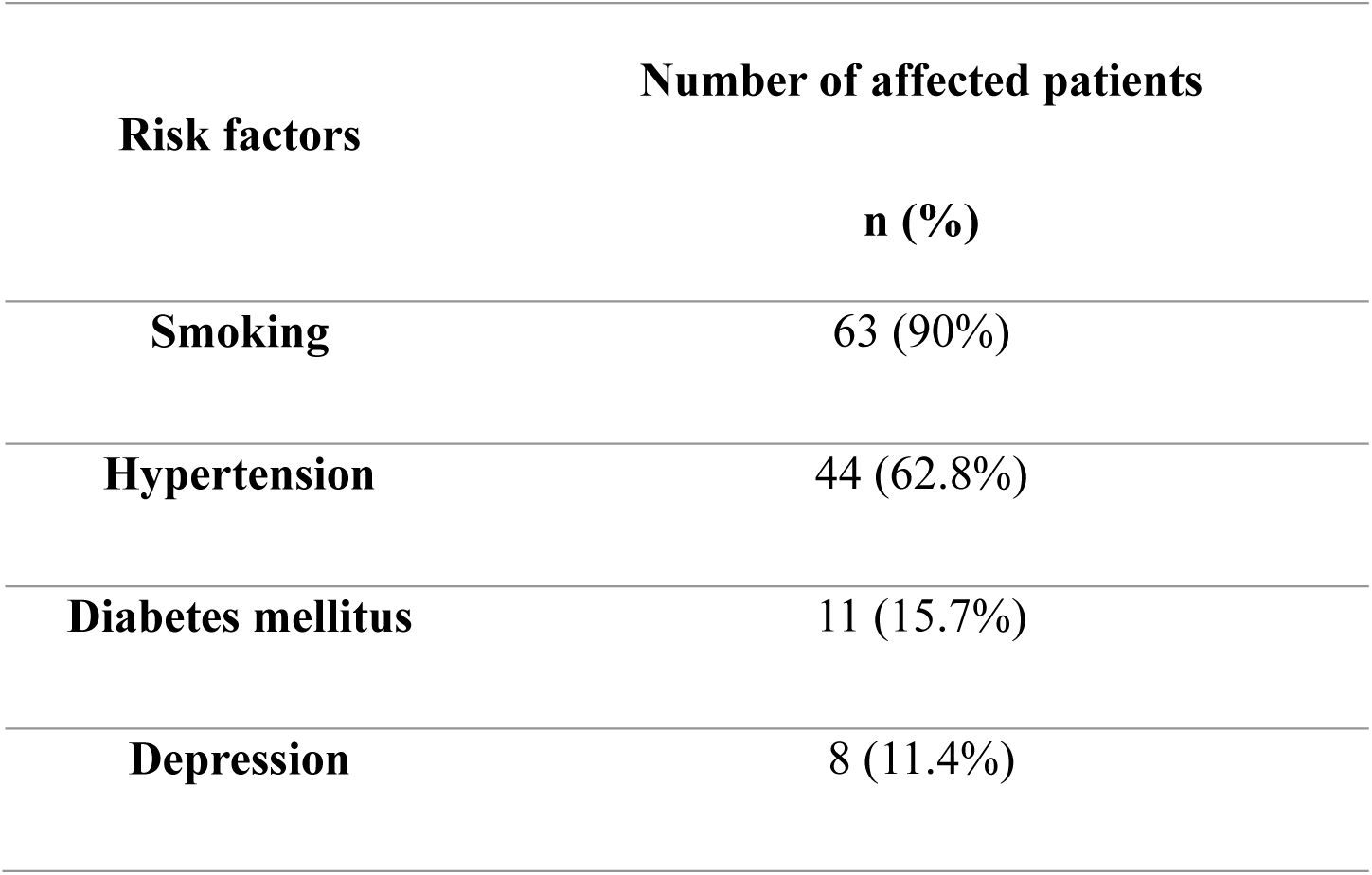
CAD risk factors in studied group (70 patients). CAD – coronary artery disease.

The mean BMI in the control group was 24.5 kg/m2, (SD ± 4.8) with 176 smokers (29,5%). The mean BMI in the control group matched with a studied group by age and gender was 26.08 kg/m2 (SD ± 4.3) and this group included 62 smokers (29,5%).

### 3.2 Next-Generation Sequencing

The sequencing encompassed the coding sequences of 159 genes with 40 nucleotide intronic flanks. Low quality variants were removed, and the variants of the allele count greater or equal to 5 in the MI group were considered in further analysis. Variants were narrowed down to those classified as at least variants of unknown significance according to the bioinformatic algorithm. This resulted in identification of four variants in *LDLR, SYNE1, PCSK9* and *APOB* genes. Allelic count of these SNPs was assessed in the control group. Furthermore, for each variant Hardy-Weinberg equilibrium was tested in the control population showing (p>0.05). Fisher’s exact test was used to check for differences in allelic frequencies in both groups; Table 4.

**Table 4.** Analyzed SNPs characteristic – number of genotypes (wild type/alternative heterozygote/alternative homozygote) in studied group (70 patients) and in control group unmatched (597 people) and matched by age and sex (210 people). A – adenine, *APOB* – apolipoprotein B gene, G – guanine, *LDLR* – low density lipoprotein receptor gene, MI – myocardial infarction, *PCSK9* – proprotein convertase subtilisin/kexin type 9 gene, SNP – single nucleotide polymorphism, *SYNE1* - spectrin repeat containing nuclear envelope protein 1 gene.

| Gene | SNP<br>(genotypes) | MI group | Control group<br>(unmatched) | Control group<br>(matched) |
| --- | --- | --- | --- | --- |
| *SYNE1* | **rs36215567**<br>(AA/AG/GG) | 64/6/0 | 586/11/0 | 208/2/0 |
| *LDLR* | **rs11669576**<br>(GG/GA/AA) | 61/7/0 | 541/48/1 | 198/10/0 |
| *PCSK9* | **rs11800265**<br>(GG/GA/AA) | 61/5/0 | 414/43/1 | 150/14/0 |
| *APOB* | **rs1801703**<br>(GG/GA/AA) | 65/5/0 | 587/9/0 | 207/3/0 |

Sequencing and statistical analysis performed in the MI patients and healthy controls revealed the presence of a single variant in the *SYNE1* gene, with an enriched abundance within the patients. We did not observe any statistically significant differences between the frequency of other variants, including known, pathogenic variants related to familial hypercholesterolemia (FH) as well as variants of unknown significance of FH related genes: *LDLR* c.667G>A, *PCSK9* c.658-36G>A, and *APOB* c.12382G>A.

*SYNE1* gene variant rs36215567 (NM_182961.4: c.20396+22A>G) is statistically significantly more prevalent in the studied group in comparison to the control population with OR 4.80, 95%CI 1.43-14.45 (p=0.005) as well as when compared to the control population matched by age and gender OR 9.31, 95%CI 1.64-95.41(p=0.004); Figure 1.

**Figure 1.**
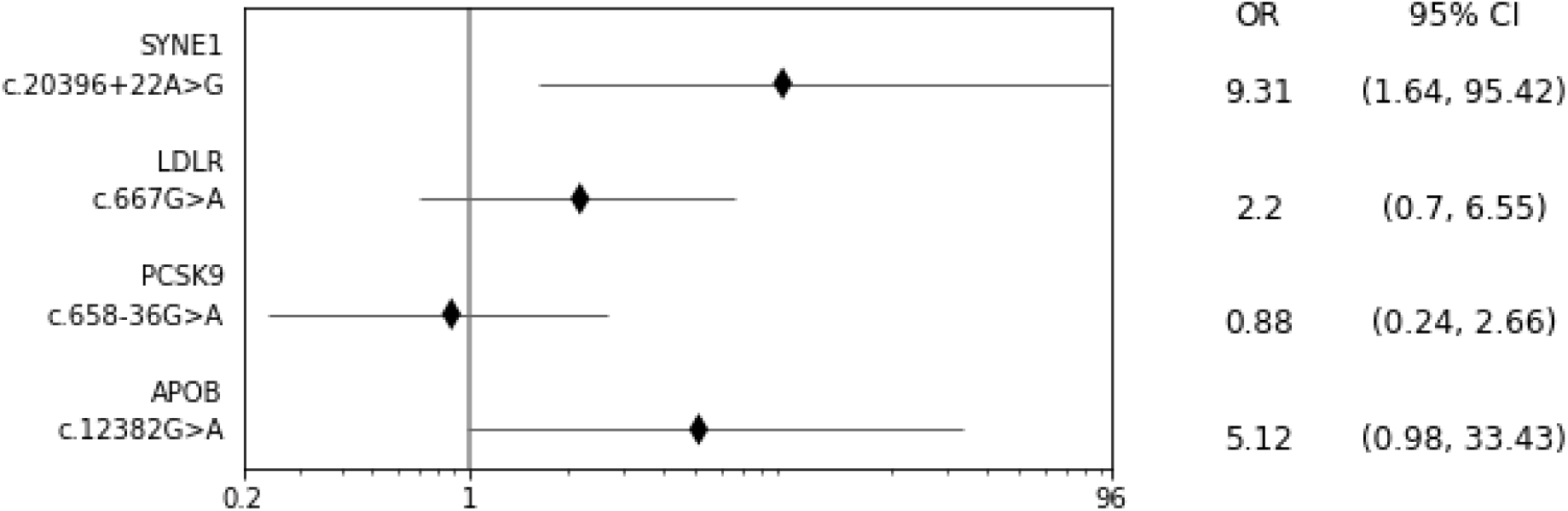
*SYNE1* gene variant rs36215567 (NM_182961.4: c.20396+22A>G) *LDLR* c.667G>A, *PCSK9* c.658-36G>A, and *APOB* c.12382G>A incidence in the studied group (70 patients) compared to the control population matched by age and gender (210 people); Fisher’s exact test. *APOB* – apolipoprotein B gene, CI – confidence intervals, G – guanine, *LDLR* – low density lipoprotein receptor gene, OR – odds ratio, *PCSK9* – proprotein convertase subtilisin/kexin type 9 gene, SNP – single nucleotide polymorphism, *SYNE1* - spectrin repeat containing nuclear envelope protein 1 gene.

Considering that the study group differed significantly in BMI and the number of smokers (p<0.001) when compared with both unmatched and matched control population the multivariable logistic regression model was created. The model included smoking status, BMI and presence of *SYNE1* rs36215567. The model confirmed this SNP to be an independent risk factor for premature MI with OR 13,86 95%CI 1.64 – 117.45 (p =0.016); Table 5.

**Table 5.** Multivariable regression model included smoking status, BMI and presence of SYNE1 rs36215567 variant confirmed this SNP as an independent risk factor for premature MI; studied group – 70 patients, control group – 597 people; Fisher’s exact test, *statistically significant. BMI – body mass index, CI – confidence intervals, MI – myocardial infarction, OR – odds ratio, SNP – single nucleotide polymorphism, SYNE1 - spectrin repeat containing nuclear envelope protein 1 gene.

| **Variable** | **OR** | **OR (95% CI)** | **P value** |
| --- | --- | --- | --- |
| **BMI** | 1.13 | 1.05 – 1.23 | 0.001* |
| **Smoking status** | 19.40 | 8.19 – 45.94 | <0.001* |
| ***SYNE1* rs36215567 variant** | 13.86 | 1.64 – 117.45 | 0.016* |

## 4. Discussion

The main aim of this study was to identify single variants predisposing to MI at a young age as the genetic background seems to be particularly important in the pathogenesis of CAD in this group. In our previous study we revealed that the younger age of patients with myocardial infarction is associated with a higher number of relatives with a history of premature MI/ischaemic stroke [12].

Numerous evidence indicates that CAD at a young age is associated with a genetic background. A significant enrichment of increased polygenic score has been noted among patients with early-onset (age ≤55 years) MI as compared with population-based controls, with median polygenic score among patients in the 72nd percentile [13]. Both familial hypercholesterolemia mutations and high polygenic score were associated with more than three-fold increased odds of early-onset MI. Monogenic familial hypercholesterolemia or a high genome-wide polygenic score confer to a 4- to 5-fold relative risk, which suggests that assessment for familial hypercholesterolemia mutations and genome-wide polygenic scores could be very useful for risk prediction, especially when combined with traditional risk factors for CAD [14].

Using data from the UK Biobank on 306,654 individuals without a history of CVD and not receiving lipid-lowering treatments, Sun L et al. calculated risk discrimination and reclassification upon addition of PRSs to risk factors in a conventional risk prediction model (i.e., age, sex, systolic blood pressure, smoking status, history of diabetes, and total and high-density lipoprotein cholesterol) [15]. The addition of information on PRSs increased the C-index by 0.012 (95% CI 0.009–0.015) and resulted in continuous net reclassification improvements – among people at intermediate (5% to <10%) 10-year CVD risk could help prevent 1 additional CVD event for approximately every 340 individuals screened. Such a targeted strategy could help prevent 7% more CVD events than conventional risk prediction alone. Moreover, it has been shown lastly, that adding PRS to clinical risk assessment has significantly improved identification people who experience a serious CVD event, especially in young age – among people aged 40-54 years clinical risk assessment alone identified 26.0% (95% CI: 16.5%–37.6%) of those who developed a major CVD event, while the combination of clinical and genetic approach identified 38.4% (95% CI: 27.2%–50.5%) [16].

In this study, we used the Next-Generation Sequencing to identify possible MI-related genetic variants in young patients. Since the familial aggregation of MI is an independent risk factor for the disease, we decided to perform an analysis using a panel of 159 genes related to various cardiovascular disorders, including 26 genes that are known to be involved in myopathies and dyslipidemias. Interestingly, our study did not show any statistically important difference in frequencies of variants commonly associated with dyslipidemias between the two studied groups [17]. We identified a single genetic variant in the *SYNE1* gene, whose prevalence was higher in MI patients. The rs36215567 (NM_182961.4: c.20396+22A>G) variant is located in an intron of a gene, possibly altering splicing of the *SYNE1* gene and resulting in altered activity of the translated protein, though the effect of this variant on the production of different *SYNE1* transcripts has not been described. It is a rare variant occurring at the frequency of 0.89% in the European population (1000 genomes project).

*SYNE1* (Spectrin Repeat Containing Nuclear Envelope Protein 1; Nesprin1) gene is a protein-coding gene located in a long arm of chromosome 6, position 6q25.2. Its protein is a member of the spectrin family of structural proteins that link the nuclear plasma membrane to the actin cytoskeleton. It is expressed in numerous tissues, but it has mainly been mapped to the aortic vascular smooth muscle cells and heart [18]. *SYNE1* aberrances were mainly associated with arthrogryposis and muscular dystrophies (Online Mendelian Inheritance in Men) but more recent papers identified its involvement in dilated cardiomyopathy [19, 20]. Interestingly, this association results mainly from the presence of alternatively spliced gene isoforms. *SYNE1* has been shown also to be involved in the angiotensin-II-induced cardiac hypertrophy by miR-525-5p / specific protein transcription factor SP1 axis [21].

Moreover, a recent study revealed an important role of *SYNE1* in cell regeneration and showed that its levels decrease with age possibly leading to a decrease in cardiac function [22]. Thus, even though the exact role of *SYNE1* protein in cardiovascular homeostasis is still largely unknown, a growing number of data suggest its role in the maintenance of proper function of the cardiac and arterial muscles.

There are also available data indicating a possible role of *SYNE1* variants (although others than shown in our study) in the development of CAD by the involvement in the morbidity of hypercholesterolemia and hypertriglyceridemia through gene-gene and SNP-SNP interactions [23]. It suggests, considering the results of our study, other potential gene-gene interactions including *SYNE1* gene variants in the pathogenesis of CAD.

The main limitation of this study is the relatively small number of participants. The assumptions of the study, including young patients with MI and a family history of premature atherosclerosis, make this population very unique, but at the same time difficult to enlarge. On the other hand, the high homogeneity of the groups, limited to the Polish Caucasian population, may be important in the context of potential population and racial differences in the pathogenesis of CAD, especially when analyzing genetic risk factors. Moreover, this the first report regarding potential role of *SYNE1* in premature CAD which means that there are no similar data in wide databases yet as well as functional studies carried out to elucidate potential mechanisms. However, this strengthens the value of our finding.

In conclusion: a novel variant of the *SYNE1* gene is associated with myocardial infarction in young age patients with a family history of premature atherosclerosis. Nevertheless, the wide use of such genetic information to predict CAD development and incidences still requires further investigation including wide genetic and functional studies before clinical implementation.

## Data Availability

All data referred to in the manuscript are available at the authors.

